# Sex Differences in the Associations between Education, Neuropathology, and Cognition

**DOI:** 10.64898/2026.09.03.26362216

**Authors:** Shima Raeesi, Yashar Zeighami, Cassandra Morrison, Mahsa Dadar

## Abstract

**IMPORTANCE:** Autopsy evidence on sex-specific associations of education with neuropathology and cognition is limited.

**OBJECTIVE:** To test sex differences in education associations with neuropathologic burden and cognition.

**DESIGN:** Cohort study of data collected from September 2005 through August 2024.

**SETTING:** Multicenter autopsy cohort from the National Alzheimer’s Coordinating Center.

**PARTICIPANTS:** Adults aged 55 years or older; cross-sectional eligibility required a final CDR-SB assessment within 2 years before death, and longitudinal eligibility additionally required at least 3 assessments.

**EXPOSURES:** Years of formal education.

**MAIN OUTCOMES AND MEASURES:** Neuropathologic measures and the Clinical Dementia Rating Scale Sum of Boxes (CDR-SB). Linear regression and mixed-effects models tested associations and interactions of education with sex, neuropathology, and linear and quadratic time. Benjamini-Hochberg correction addressed multiple comparisons.

**RESULTS:** Among 2592 participants (mean [SD] age at death, 81.6 [10.8] years; 1188 [45.8%] female), 1969 contributed to longitudinal analyses. In female participants, higher education was associated with lower Thal amyloid phase, diffuse plaques, neuritic plaques, and Braak stage (β range, −0.097 to −0.071; *q_FDR_*<.026); none was significant among male participants. Education-by-sex interactions supported differences in Thal amyloid phase, and diffuse and neuritic plaques (β range, −0.119 to −0.097; *q_FDR_*<.045). Education was unrelated to final CDR-SB among female (β, −0.007; 95% CI, −0.052 to 0.038; *P*=.767) and male participants (β, −0.026; 95% CI, −0.071 to 0.019; *P*=.254), with no education-by-sex interaction (*P*=.660). The education-by-Braak-stage-by-sex interaction indicated that higher education was associated with a weaker Braak stage-CDR-SB association among females than males (β, −0.104; 95% CI, −0.171 to −0.038; *q_FDR_*=.022). Longitudinally, education was unrelated to linear CDR-SB change in either sex. Education-by-quadratic-time associations were observed among females (β, 0.002; 95% CI, 0.001-0.003; *P*<.001) and male participants (β, 0.001; 95% CI, 0-0.002; *P*=.037). The corresponding interaction with sex was also significant (*P*=.043).

**CONCLUSIONS AND RELEVANCE:** Among female participants, higher education was associated with lower selected Alzheimer’s disease neuropathologic measures and weaker associations of tau pathology with cognition. Longitudinal trajectories were nonlinear, with slower earlier decline followed by later acceleration at higher education levels.

**Key Points:** *Question:* Do associations between education, neuropathologic burden and cognition differ by sex?

*Findings:* In this cohort study of 2592 autopsy participants, higher education was associated with lower Alzheimer’s disease neuropathologic burden among female but not male participants, and education-by-sex interactions were significant for Thal amyloid phase, diffuse and neuritic plaques. Higher education was associated with a weaker Braak stage-CDR-SB association among female than male participants and with lower CDR-SB five years before death in both sexes. The associations persisted after adjustment for neuropathologic burden.

*Meaning:* Education-neuropathology associations and the clinical expression of neuropathology may be sex-specific.

## Introduction

The cognitive consequences of neuropathologic burden vary markedly across individuals in Alzheimer’s disease (AD) and related dementias^1–7^: some individuals show increasingly deteriorating cognition as pathology accumulates, whereas others maintain better-than-expected cognition despite comparable neuropathologic burden^3–9^, a pattern commonly explained through cognitive reserve. Autopsy studies suggest that educational attainment may be associated with neuropathologic burden and may modify the association between neuropathology and cognition^6,7,10–13^; however, evidence is inconsistent across neuropathologic measures and analytic frameworks^1,8,10,12,14,15^. Potential sex differences in these associations may reflect biological variation in susceptibility to neuropathology and gendered variation in educational opportunities^1,4,5,8,16^. Prior studies have often focused on a limited set of predominantly AD-related pathologies, used a single cognitive assessment timepoint near death, or presented sex-stratified estimates without formal interaction testing^1,2,5,7,10,11,16^. Consequently, it remains unclear whether associations of education with cross-sectional and longitudinal cognition differ by sex in the presence of multiple coexisting pathologies.

Using the National Alzheimer’s Coordinating Center (NACC)^17,18^ autopsy cohort, this study tested prespecified hypotheses that associations of education with neuropathologic measures, final Clinical Dementia Rating Scale Sum of Boxes (CDR-SB) score^19^, and predeath cognitive trajectories would differ by sex. Robustness was evaluated in age-, neuropathology-, and covariate-matched samples.

## Methods

### Study Cohort

This retrospective multicenter autopsy cohort study used clinical and postmortem assessments from the NACC Uniform and Neuropathology Data Set^17,18^ (data freeze 67, September 2024). Written informed consent was obtained and documented at each contributing National Institute on Aging-AD Research Center (ADRC) under institutional review board-approved protocols before data submission to NACC. Participants aged 55 years or older at death were included to focus on later-life neuropathologic and cognitive-functional changes. Years of formal education were analyzed continuously. Autopsies and neuropathologic assessments were performed at contributing ADRCs using standardized NACC Neuropathology Data Forms^17^. Neuropathologic measures representing AD and non-AD processes were examined. The primary cognitive-functional outcome was the CDR-SB (range, 0-18)^19^, with higher scores indicating greater impairment. Cross-sectional analyses used the last valid CDR-SB assessment within 2 years before death.

Longitudinal analyses included participants with at least 3 valid predeath assessments and a final assessment within this interval (eFigure 1). Prespecified covariates were age at death, race and ethnicity, APOE ε4 carrier status, and postmortem interval. Sex was included in the combined-sex interaction models testing between-sex differences. Cross-sectional models additionally included the final assessment-to-death interval, whereas longitudinal models represented assessment timing as time relative to death. APOE ε4 carrier status indicated the presence of at least one ε4 allele. Invalid, unknown, or unassessed values were excluded. Continuous variables and covariates were standardized using means and standard deviations from the corresponding combined-sex analytic sample before sex stratification. Participant numbers and demographic characteristics are reported in the Results.

### Statistical Analysis

Participant characteristics were summarized by sex and compared using Welch 2-sample t tests, Wilcoxon rank-sum tests, and Pearson χ² or Fisher exact tests, as appropriate. Sex-stratified associations of education with neuropathologic measures were estimated using linear regression for ordinal outcomes and logistic regression for binary outcomes. Education-by-sex interaction models formally tested between-sex differences. Within each sex, linear regression related education to final CDR-SB before and after neuropathologic adjustment, while education-by-pathology interactions evaluated whether education modified the association of each neuropathologic measure with CDR-SB. Combined-sex education-by-pathology-by-sex interactions compared this modification between sexes. Each interaction model included the other neuropathologic measures and all corresponding main effects and lower-order interactions.

Predeath CDR-SB trajectories were modeled using linear mixed-effects models with participant-specific random intercepts and linear time slopes. Time was expressed in years relative to death and centered at the mean assessment time, approximately 5.1 years before death. Linear models included education-by-time interactions; quadratic models additionally included time² and education-by-time² interactions. Sex-stratified and combined-sex interaction models were fitted with and without neuropathologic adjustment. Mixed-effects models used maximum likelihood, and likelihood-ratio tests compared linear and quadratic fits. Benjamini-Hochberg false-discovery rate (FDR) correction^20^ was applied separately within each prespecified family of neuropathology or interaction tests, with *q_FDR_*<.05 considered statistically significant. For other analyses, *P*<.05 was considered statistically significant. Regression coefficients for linear and mixed-effects models and odds ratios for logistic models are reported with 95% confidence intervals (CIs). Analyses were conducted using *R*, version 4.1.3 (*R Foundation for Statistical Computing*).

### Sensitivity Analyses

Three matching strategies addressed age differences between sexes and neuropathologic or covariate differences between education groups. Female and male participants were matched 1:1 on age at death to assess whether age differences accounted for the sex-specific findings. To compare education-related cognitive-functional outcomes among participants with similar neuropathologic burden, lower-education participants (<16 years) were matched 1:1 to higher-education participants (≥16 years), once on neuropathologic measures alone and once on these measures plus measured covariates. The education threshold, based on the sample median, was used only to construct the matched samples; all postmatch outcome models analyzed education continuously. Additional matching procedures, postmatch analysis details, and diagnostics are provided in the eMethods.

## Results

### Cohort Characteristics

Among 2592 participants, 1188 (45.8%) were female and 1404 (54.2%) were male. Most participants were non-Hispanic White (2379 [91.8%]), and 1080 (41.7%) were APOE ε4 carriers. Female participants were older at death than male participants (mean [SD], 83.2 [10.9] vs 80.2 [10.5] years) and had fewer years of education (15.1 [2.7] vs 16.5 [3.0] years; *P*<.001 for both). Race and ethnicity also differed by sex (*P*=.004). After FDR correction, the distributions of neuritic plaques, Braak stage, Lewy body pathology, cerebral cortical atrophy, and hippocampal atrophy differed by sex (Table 1).

**Table 1.**
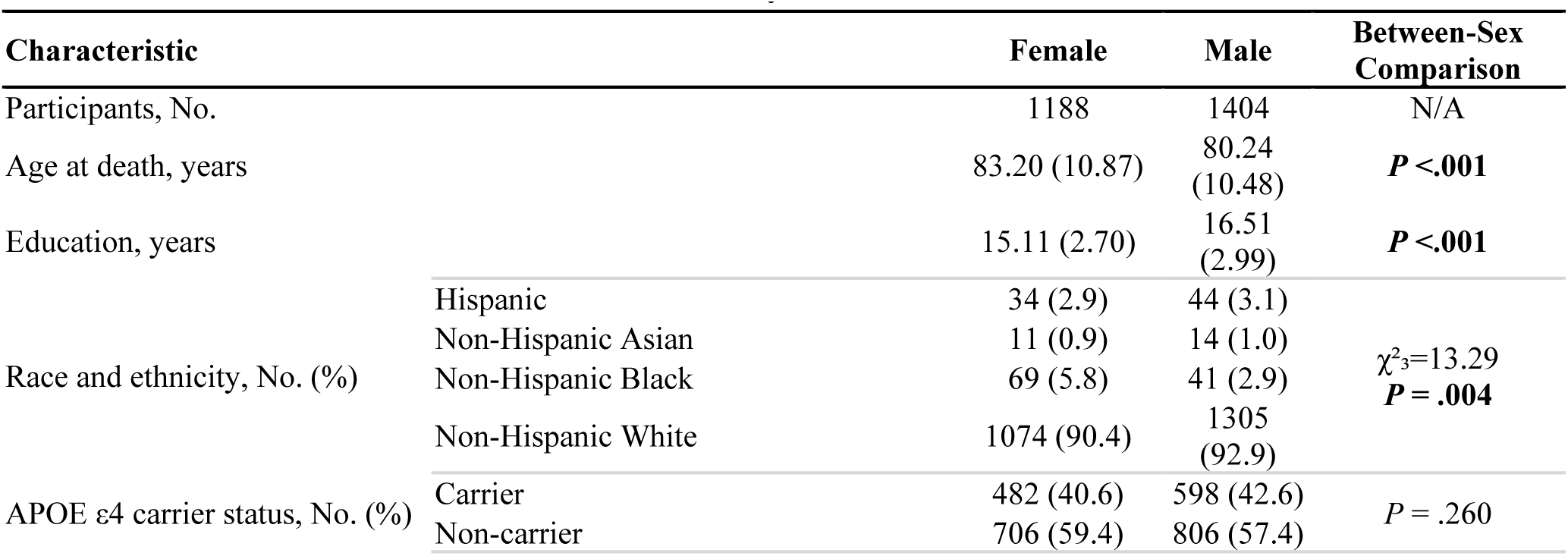
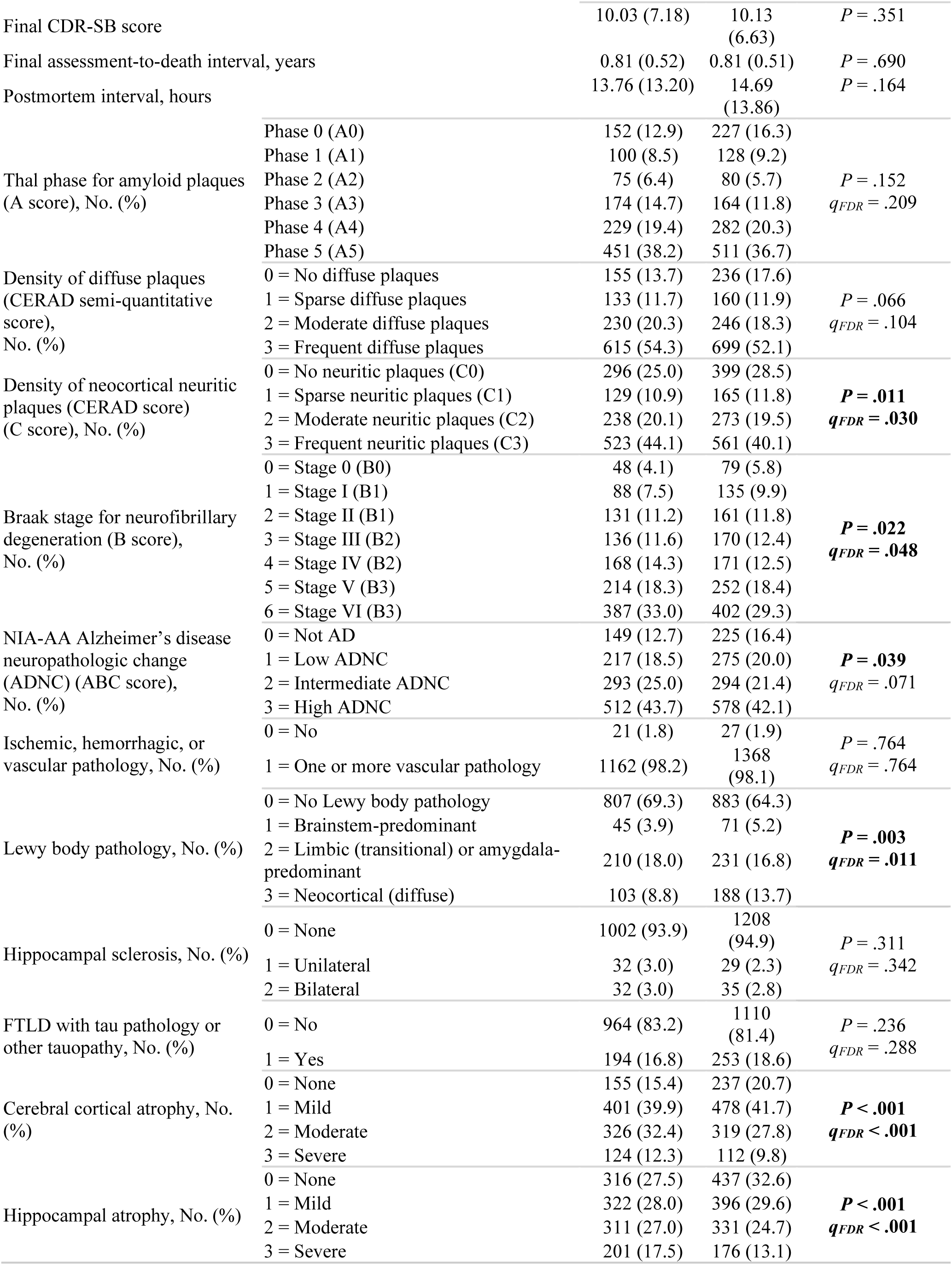
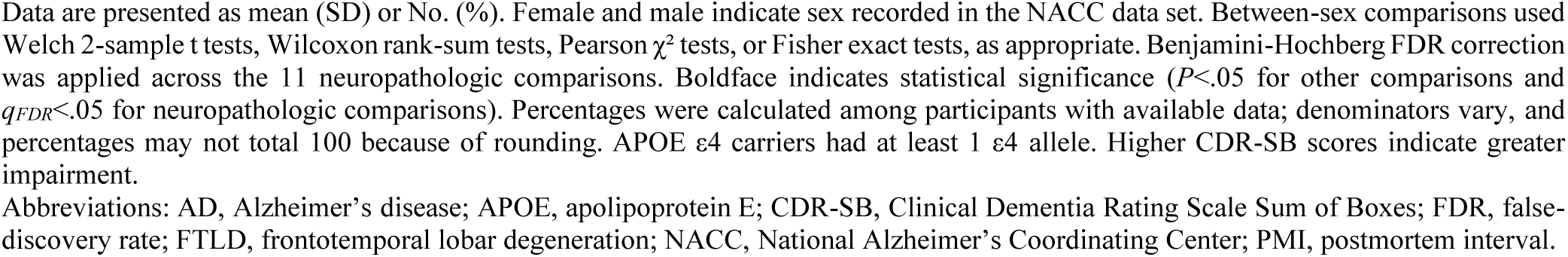
Characteristics of the Source Cohort by Sex.

### Education and Neuropathologic Measures

Higher education was associated with lower Thal amyloid phase, diffuse plaques, neuritic plaques, Braak stage, and AD neuropathologic change among female participants (all *q_FDR_*<.05), whereas no associations survived FDR correction among males (Table 2, eFigure 2). The sex-stratified estimates were similar after female and male participants were matched on age at death (eTable 1). Education-by-sex interactions were significant for Thal amyloid phase (β, −0.110; 95% CI, −0.185 to −0.035; *q_FDR_*=.022), diffuse plaques (β, −0.119; 95% CI, −0.197 to −0.042; *q_FDR_*=.022), and neuritic plaques (β, −0.097; 95% CI, −0.172 to −0.021; *q_FDR_*=.045; eTable 2, Figure 1).

**Figure 1.**
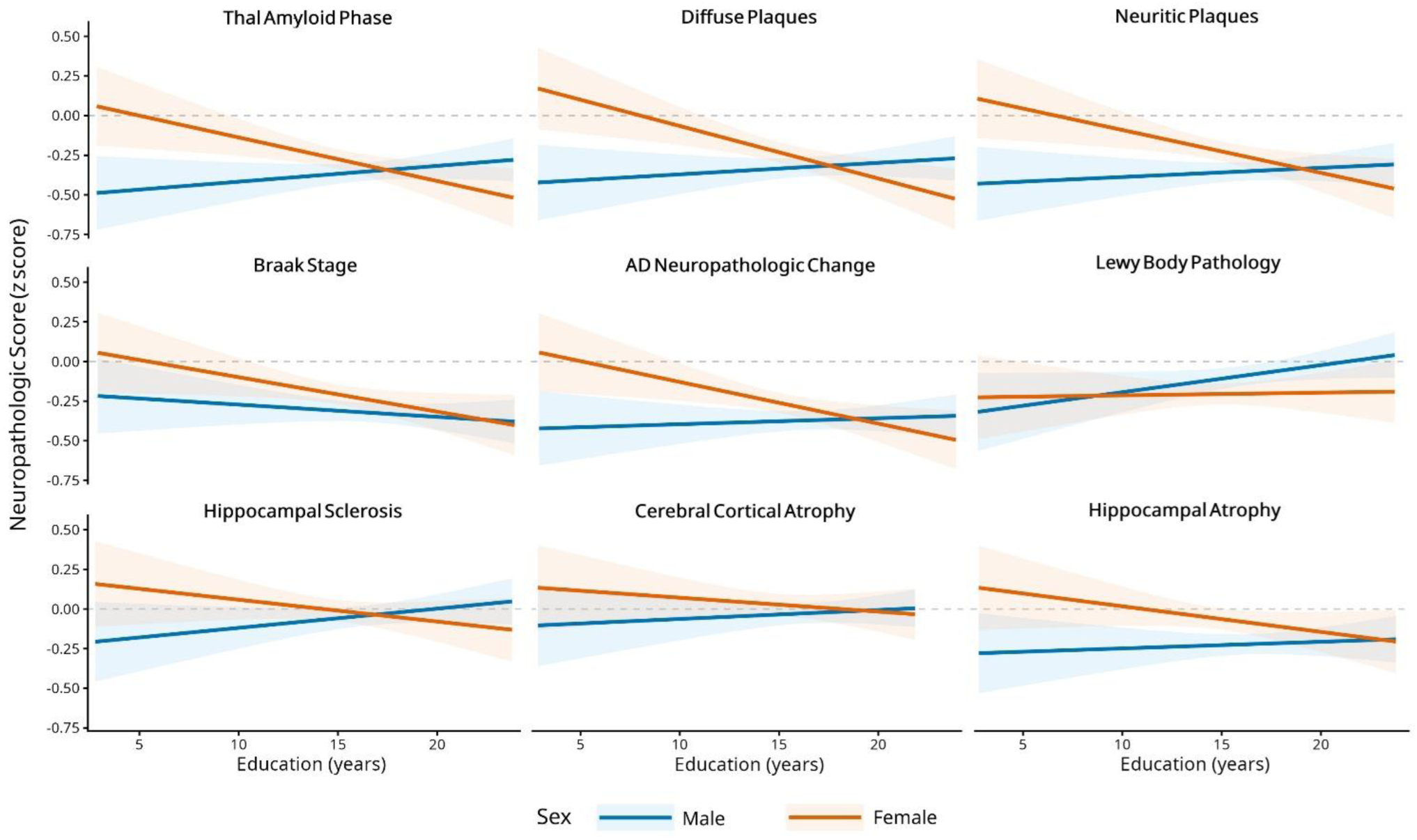
Sex-Specific Associations Between Education and Neuropathologic Measures. Each panel shows model-predicted scores for one standardized ordinal neuropathologic measure across years of education among female and male participants. A separate multivariable linear regression model tested the education-by-sex interaction for each measure using the following covariate structure: ***Neuropathology ∼ Education × Sex + Age at death + APOE ε4 carrier status + Race and ethnicity + PMI*** All corresponding main effects were retained. Male participants were the reference group. Lines represent model-predicted values, and shaded areas represent 95% CIs. Education, ordinal neuropathologic measures, and continuous covariates were standardized using means and SDs from the corresponding combined-sex analytic sample; education is displayed in original years. Predictions were limited to the observed education range for each analytic sample, with continuous covariates held at their means, race and ethnicity set to non-Hispanic White, and APOE ε4 carrier status set to noncarrier. Lower z scores indicate lower levels of the corresponding neuropathologic measure. Benjamini-Hochberg FDR correction was applied across the education-by-sex interaction tests. Binary measures (vascular pathology and FTLD-tau or other tauopathy) were analyzed using logistic regression and were not displayed because their predicted probabilities are not directly comparable with standardized ordinal outcomes. Female and male indicate sex recorded in the NACC data set. Abbreviations: AD, Alzheimer’s disease; APOE, apolipoprotein E; CI, confidence interval; FDR, false-discovery rate; FTLD, frontotemporal lobar degeneration; NACC, National Alzheimer’s Coordinating Center; PMI, postmortem interval; SD, standard deviation.

**Table 2.**
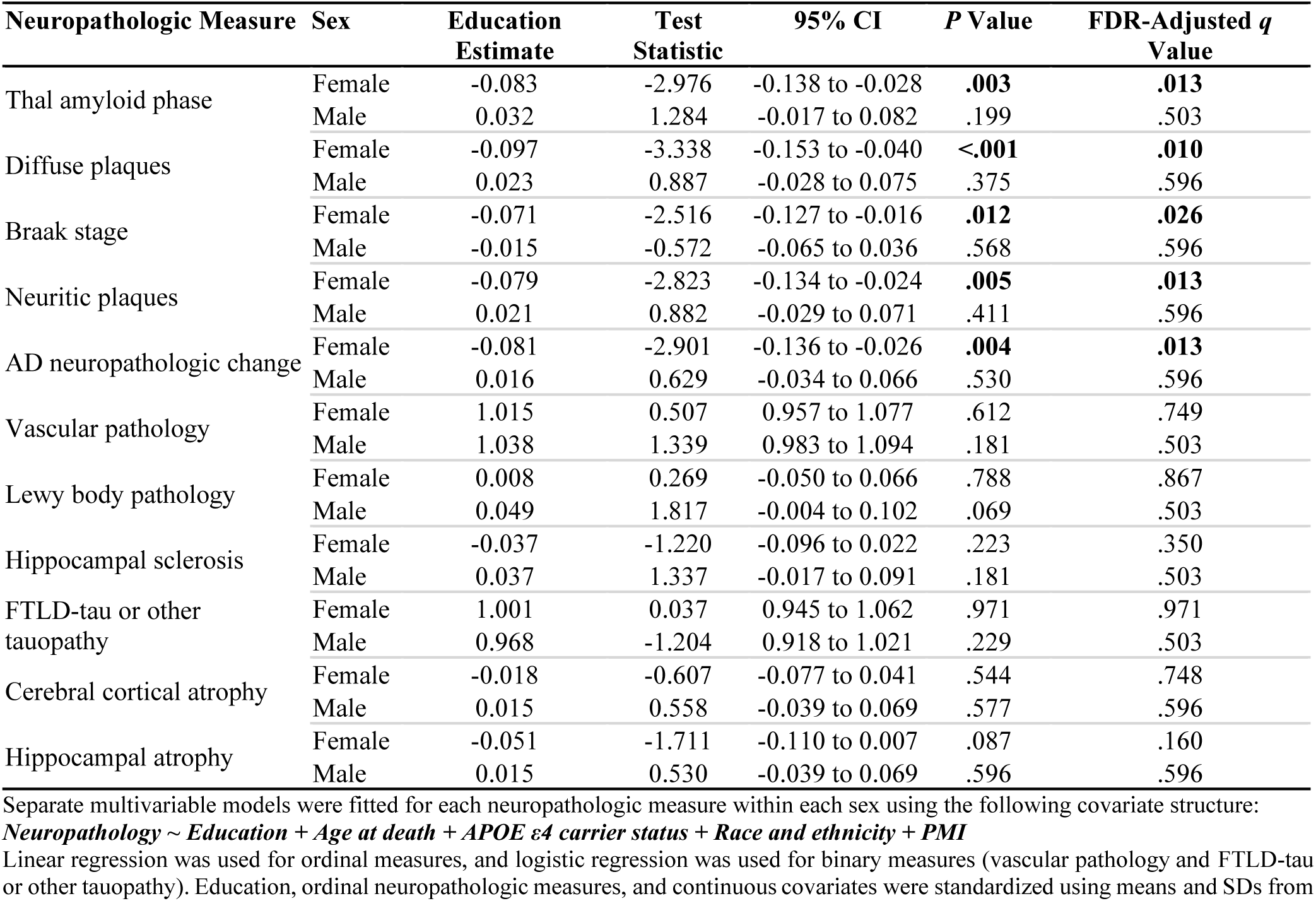
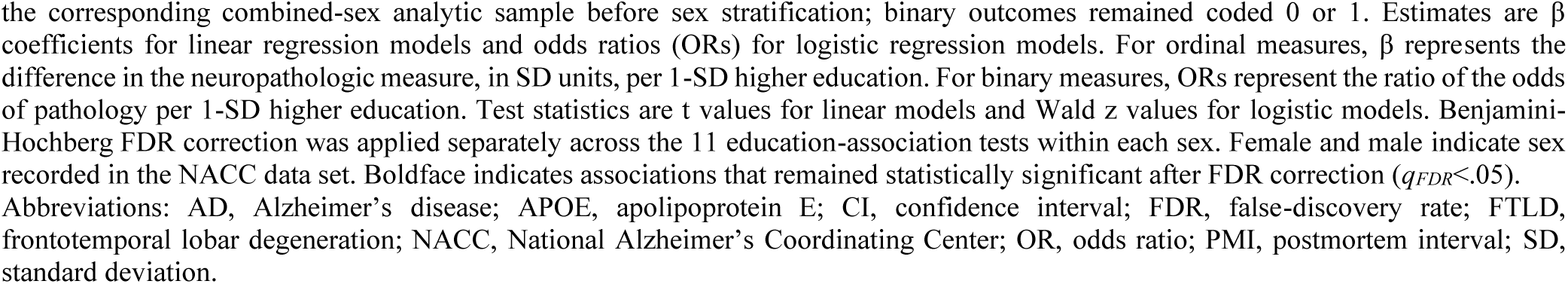
Sex-Stratified Associations Between Education and Neuropathologic Measures.

### Cross-sectional Cognition Associations

After adjustment for all neuropathologic measures, education was not associated with final CDR-SB among female (β, −0.007; 95% CI, −0.052 to 0.038; *P*=.767) or male participants (β, −0.026; 95% CI, −0.071 to 0.019; *P*=.254). The education-by-sex interaction was also not statistically significant (β, 0.015; 95% CI, −0.050 to 0.079; *P*=.660). These null associations persisted across all matched analyses (eTables 3 and 4). Among female participants, higher education was associated with weaker associations of neuritic plaques (β, −0.077; 95% CI, −0.127 to −0.028; *q_FDR_*=.013) and Braak stage (β, −0.082; 95% CI, - 0.134 to −0.029; *q_FDR_*=.013) with CDR-SB; neither interaction was statistically significant among male participants (eTable 5 and Figure 2). The education-by-Braak-stage-by-sex interaction survived FDR correction (β, −0.104; 95% CI, −0.171 to −0.038; *q_FDR_*=.022). After age matching, the education-by-Braak-stage and education-by-neuritic-plaque interactions remained significant among female participants (eTables 6 and 7).

**Figure 2.**
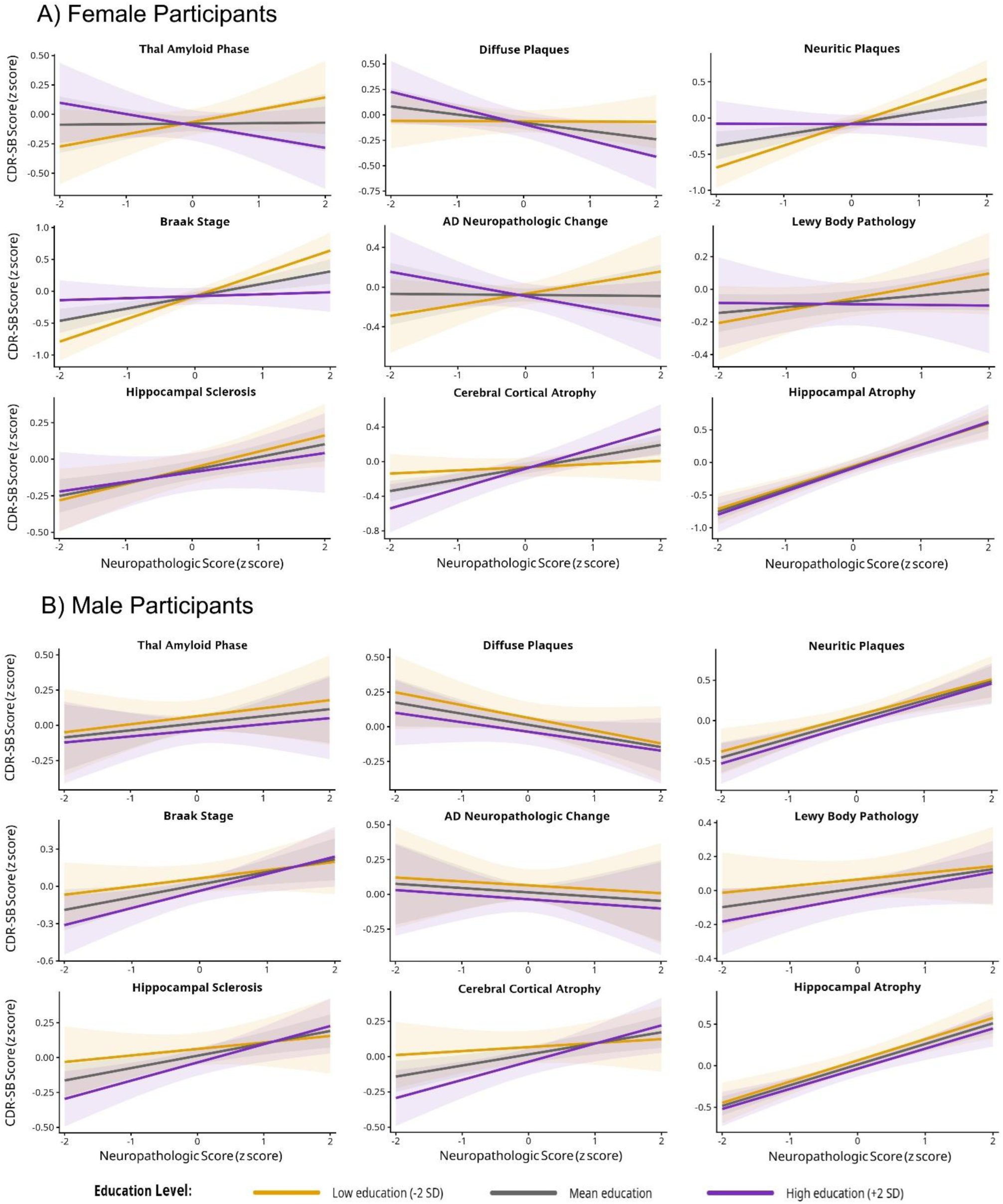
Associations Between Neuropathologic Measures and Final CDR-SB at Selected Education Levels by Sex. Panel A shows model-predicted standardized final CDR-SB scores among female participants, and panel B shows the corresponding predictions among male participants. Lines represent education values 2 SDs below the mean, at the mean, and 2 SDs above the mean of the corresponding combined-sex analytic sample. These values were selected for visualization; education was analyzed continuously. For each focal ordinal neuropathologic measure, predictions were generated from separate sex-stratified multivariable linear regression models using the following covariate structure: ***Final CDR-SB ∼ Education × Neuropathology + Age at death + APOE ε4 carrier status + Race and ethnicity + PMI + Final assessment-to-death interval + Other neuropathologic measures*** All corresponding main effects and lower-order interactions were retained. Formal between-sex differences were tested using corresponding combined-sex education-by-neuropathology-by-sex interaction models, with male participants as the reference group. Education, final CDR-SB, ordinal neuropathologic measures, and continuous covariates were standardized using means and SDs from the corresponding combined-sex analytic sample before sex stratification. Predictions were restricted to the observed standardized range of each focal neuropathologic measure, with continuous covariates and the other neuropathologic measures held at their means, race and ethnicity set to non-Hispanic White, and APOE ε4 carrier status set to noncarrier. Shaded areas represent 95% CIs. Benjamini-Hochberg FDR correction was applied separately across the education-by-neuropathology interaction tests within each sex and across the education-by-neuropathology-by-sex interaction tests. Higher CDR-SB scores indicate greater cognitive and functional impairment. Binary neuropathologic measures (vascular pathology and FTLD-tau or other tauopathy) were included as 0/1 predictors in the same linear regression framework but were not displayed because the continuous standardized neuropathology axis used for visualization is not meaningful for binary variables. Female and male indicate sex recorded in the NACC data set. Abbreviations: AD, Alzheimer’s disease; APOE, apolipoprotein E; CDR-SB, Clinical Dementia Rating Scale Sum of Boxes; CI, confidence interval; FDR, false-discovery rate; FTLD, frontotemporal lobar degeneration; NACC, National Alzheimer’s Coordinating Center; PMI, postmortem interval; SD, standard deviation.

### Longitudinal CDR-SB Trajectories

The longitudinal sample included 1969 participants contributing 14246 observations (mean, 7.2 per participant), spanning up to 17.9 years before death. In neuropathology-adjusted sex-specific models, higher education was associated with lower predicted CDR-SB at the centered time point, approximately five years before death, among females (β, −0.054; 95% CI, −0.099 to −0.009; *P*=.018) and male participants (β, −0.053; 95% CI, −0.090 to −0.015; *P*=.006). Education-by-time interactions were not significant in either sex (both *P*>.05). The quadratic model provided a better fit than the linear model (likelihood-ratio χ²₂=873.63; *P*<.001). Education-by-time² interactions were significant in both sexes in the primary and age-matched samples but only among female participants in the 2 education-group-matched samples. The education-by-time²-by-sex interaction was also significant in the primary sample (β, 0.001; 95% CI, 0.000-0.002; *P*=.043) and the neuropathology- and covariate-matched sample (*P*=.004), but not in the age-matched or neuropathology-matched samples. Thus, evidence of sex differences in nonlinear education-related trajectories was inconsistent across samples (eTables 8-11, Figure 3, eFigure 3). In both sexes, adding neuropathologic measures as covariates increased the variance explained by the models, as indicated by higher adjusted R² in cross-sectional analyses and higher marginal R² in longitudinal analyses.

**Figure 3.**
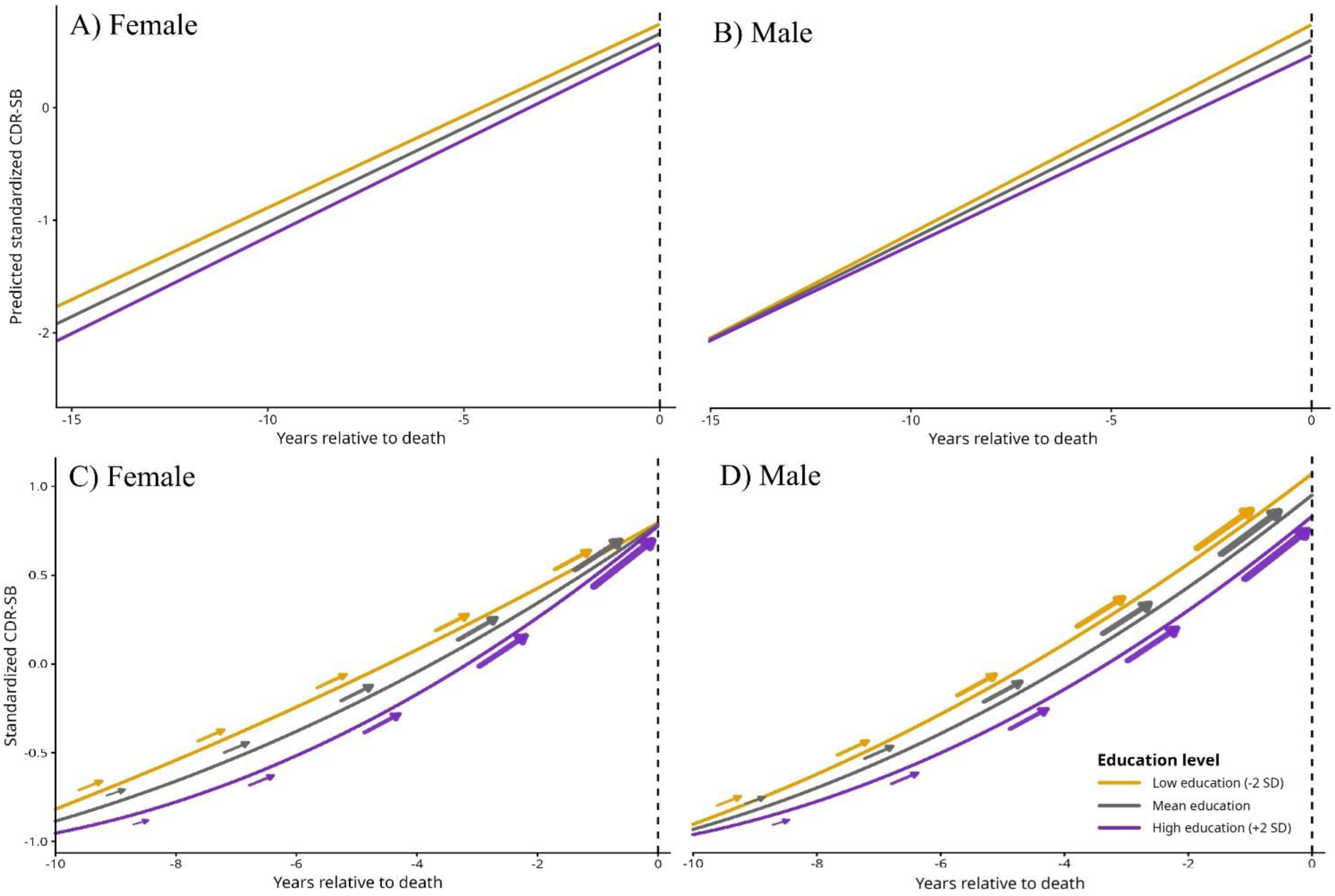
Linear and Quadratic Predeath CDR-SB Trajectories at Representative Education Levels by Sex. The primary sample refers to the original unmatched longitudinal analytic sample. Panels A and B show model-predicted linear CDR-SB trajectories for female and male participants, respectively; panels C and D show the corresponding quadratic trajectories. Predictions are presented at education values 2 SDs below the mean, at the mean, and 2 SDs above the mean. These values were selected for visualization; education was analyzed continuously. Neuropathology-adjusted models were fit separately for female and male participants: ᵃ Linear model: ***CDR-SB ∼ Education × Time + Age at death + APOE ε4 carrier status + Race and ethnicity + PMI + Neuropathologic measures + (1 + Time* ∣ *Participant ID)*** ᵇ Quadratic model: ***CDR-SB ∼ Education × (Time + Time ²) + Age at death + APOE ε4 carrier status + Race and ethnicity + PMI + Neuropathologic measures + (1 + Time* ∣ *Participant ID)*** All corresponding main effects and lower-order interaction terms were retained. Female and male refer to sex recorded in the NACC data set. Time was expressed in years relative to death, with death defined as 0, and centered at the mean assessment time, approximately 5.1 years before death. Time² was calculated from centered time, and time was not standardized. Continuous variables other than time were standardized using means and SDs from the corresponding combined-sex analytic sample before sex stratification. Trajectories are displayed using the original time scale. Model-predicted values were calculated with continuous covariates and neuropathologic measures held at their means, race and ethnicity set to non-Hispanic White, and APOE ε4 carrier status set to noncarrier. In the quadratic panels, arrows represent the instantaneous model-estimated direction and rate of CDR-SB change along each trajectory. The vertical dashed line indicates death. Higher CDR-SB scores indicate greater cognitive and functional impairment. Abbreviations: APOE, apolipoprotein E; CDR-SB, Clinical Dementia Rating Scale Sum of Boxes; NACC, National Alzheimer’s Coordinating Center; PMI, postmortem interval.

## Discussion

The present study provides autopsy-based evidence that education-pathology and education-cognition associations may be sex-dependent. Higher education was associated with lower Thal amyloid phase, diffuse and neuritic plaque density, and Braak stage among female participants, with formal interactions supporting sex differences for Thal amyloid phase, diffuse plaques, and neuritic plaques. Education was not directly associated with final CDR-SB in either sex. Among female participants, higher education was associated with weaker associations of neuritic plaques and Braak stage with CDR-SB, although only the education-by-Braak-stage interaction differed significantly by sex. Longitudinally, higher education was associated with lower CDR-SB approximately five years before death in both sexes, but evidence that the nonlinear association differed by sex was inconsistent across matched analyses. The inverse education-pathology associations among female participants align with a prior combined-sex autopsy study reporting lower AD neuropathologic burden among individuals with higher education^21^. Related autopsy studies linked greater early-life cognitive enrichment^22^ and stronger linguistic ability in young adulthood^23^ to lower global AD pathology and fewer neocortical neurofibrillary tangles, respectively, although these exposures were broader than educational attainment alone. Amyloid-positron emission tomography (PET) studies similarly linked higher educational attainment to lower cortical amyloid burden^24–27^, although a meta-analysis identified a stage-dependent association^28^. However, the broader evidence is inconsistent: several autopsy cohorts found little or no association between education and AD or other neuropathologic markers^21,29–31^ and some imaging studies were also null^24,32,33^.

The present findings extend this literature by showing inverse education-pathology associations among female participants, with formal interactions supporting sex differences for amyloid measures. Analyses combining sexes may therefore partly obscure sex-specific associations, although these observational findings do not establish that education reduces AD pathology.

The education-by-Braak-stage-by-sex interaction provided the clearest evidence that the relationship between education, neuropathology, and cognitive-functional impairment differed by sex. Among female participants, higher education was associated with weaker Braak stage- and neuritic plaque density-CDR-SB associations, suggesting less cognitive impairment at a given level of pathology; however, only the Braak-stage interaction differed significantly by sex.

While some prior studies associated higher education with better cognition proximate to death^11,34,35^, others found narrowing education-related cognitive differences with advancing AD pathology^36,37^. Evidence for modification of neuropathology-cognition associations has also been mixed, with reports of modest moderations on the associations between amyloid and neuritic plaques and cognition^38,39^, or no associations^29^, and findings for neurofibrillary tangles have been less consistent^11,35,40,38^. Finally, the postmortem literature supports the notion that the cognitive consequences of neuropathology may differ between females and males^40–42^. Similarly, an in-vivo longitudinal imaging study reported sex differences in the extent to which education modified the cortical thickness-cognition associations^43^.

Within this mixed literature, the present findings support a sex-specific difference in the cognitive expression of Braak stage rather than generalized cognitive reserve among female participants. Notably, the neuropathologic measures associated with education partly differed according to whether neuropathologic burden or CDR-SB was modeled as the outcome^11,29^. Because Braak staging captures the distribution of neurofibrillary tau pathology only at autopsy, it does not represent within-person progression through successive stages, and these analyses cannot determine when education-related cognitive differences emerged.

Although previous studies document sex differences in AD neuropathology and cognition, they do not explain why inverse education-pathology associations emerged primarily among female participants^5,16,40–44^. Greater clinical consequences of AD pathology and greater tau burden or accumulation have been reported among females^5,16,41,42,45,46^, but these findings do not establish a mechanism linking education to lower neuropathologic burden. Social and historical selection may also contribute^47–50^. The same years of schooling may represent different educational quality, occupational opportunities, and socioeconomic conditions across sexes and birth cohorts, and females who attained higher education may have constituted a particularly selected group^1,4,8,47–49,51^. Persistence of the main findings after age matching suggests that they were not explained solely by age differences between female and male participants. Unfortunately, educational quality, literacy, occupation, gender-related exposures, reproductive factors, differential survival, and brain-donation selection could not be evaluated in this cohort. The observed pattern may therefore reflect a combination of biological vulnerability, socially patterned educational opportunity, cohort and selection effects, or residual confounding, rather than education altering AD pathogenesis differently by sex.

The absence of a direct association between education and final CDR-SB in either sex contrasts with previous studies associating more education with better cognitive performance proximate to death^34^ and lower CDR-SB^34,52^. Differences in assessment timing and outcome measurement may partly explain this discrepancy. Final assessments occurred close to death, when advanced disease and convergence of cognitive trajectories may attenuate education-related differences in both sexes^12,53^. Higher education was associated with lower CDR-SB approximately five years before death in both sexes and was unrelated to linear CDR-SB change. Previous studies have similarly associated education with better cognitive performance, but not subsequent change^51,54–56^, although slower decline has been reported in specific cognitive domains^57–59^. Longitudinal studies using CDR-SB and related outcomes found faster decline among females with mild cognitive impairment and the slowest decline among highly educated males^60,61^; these sex differences were absent among participants with AD dementia^60–63^. These contrasting findings suggest that sex differences in education-related cognitive trajectories may vary by clinical stage, cognitive outcome, and cohort composition. Their attenuation in later disease stages is consistent with the similar final CDR-SB findings among female and male participants in the present study.

Education was associated with quadratic CDR-SB change, consistent with findings linking higher education to a later onset of accelerated or terminal decline followed by a shorter period of rapid deterioration^53,64–66^. Higher cognitive reserve has similarly been related to better cognitive performance before impairment but faster subsequent decline^37,67–69^. The quadratic association was observed in both sexes in the primary analyses and persisted across matched samples among female participants, whereas findings among male participants were inconsistent. The education-by-quadratic-time-by-sex interaction was also inconsistent across matched samples, providing no evidence of a robust sex-specific nonlinear effect. Moreover, quadratic models characterize nonlinear change without identifying when acceleration begins and therefore cannot establish that education delayed decline. Overall, sex differences were more evident for education-neuropathology and neuropathology-CDR-SB associations than for direct education-CDR-SB associations or trajectories. This pattern suggests that sex may be more relevant to how education relates to neuropathologic burden and its cognitive expression than to direct education-related differences in cognitive decline.

### Strengths and Limitations

This study has several strengths, including a large multicenter cohort, standardized neuropathologic data, formal testing of sex interactions, and complementary cross-sectional, longitudinal, and matched analyses. Several limitations should be acknowledged. The referral-, volunteer-, and autopsy-based composition of NACC may preferentially include more highly educated participants and individuals who survived long enough and consented to brain donation, potentially introducing selection and survival biases. Sex was recorded as a binary variable without information on gender identity or gender-related exposures. Educational attainment, measured as years of education, is a limited proxy for cognitive reserve and does not capture educational quality, literacy, occupation, or leisure and social activities, which may themselves differ by sex^4,48–50^. The predominantly non-Hispanic White sample limits generalizability and the examination of intersecting sex, racial, ethnic, and socioeconomic factors. The female-specific findings therefore require replication in independent autopsy cohorts. Neuropathologic measures were semiquantitative ordinal categories; therefore, the estimates represent modeled trends across recorded categories rather than continuous biological dose-response relationships. Neuropathology was assessed at autopsy, whereas CDR-SB was assessed before death, resulting in temporal mismatch despite restricting final assessments to within 2 years of death. Matching improved balance in measured characteristics but could not eliminate residual confounding or establish causality; matching on neuropathology may also have conditioned on a factor downstream of education. The absence of a direct education association with final CDR-SB does not exclude earlier education-related cognitive differences that diminished closer to death. Finally, the quadratic models imposed smooth trajectories, did not identify when acceleration began, and may not capture discrete change points or heterogeneity in terminal decline.

## Conclusions

This study provides postmortem evidence that selected associations of education with AD neuropathologic burden and with the neuropathology-cognition relationship may differ by sex. Higher education was associated with lower AD neuropathologic burden among female participants, with formal sex differences for Thal amyloid phase, diffuse plaques, and neuritic plaques. Higher education was also associated with weaker Braak stage-CDR-SB and neuritic plaque-CDR-SB associations in female participants, although only the education-by-Braak-stage-by-sex interaction formally supported a sex difference. These findings suggest that education may be related to both neuropathologic burden and the clinical expression of pathology among female participants, although causal effects on pathologic accumulation cannot be inferred. Pooled analyses may obscure female-specific associations.

## Data Availability

The deidentified participant-level data used in this study are available from the National Alzheimer's Coordinating Center through its Data Request Process, subject to approval of the request and completion of the National Alzheimer's Coordinating Center Data Use Agreement. The authors do not own these data and are not authorized to redistribute them.

https://www.naccdata.org/data-request-process/

## Acknowledgment

Data were obtained from the National Alzheimer’s Coordinating Center (NACC). The authors thank the participants who donated their brains, their families, and the clinicians and researchers who collected the clinical and neuropathologic data. The NACC database is funded by NIA/NIH Grant U24 AG072122. NACC data are contributed by the NIA-funded ADRCs: P30 AG062429 (PI James Brewer, MD, PhD), P30 AG066468 (PI Oscar Lopez, MD), P30 AG062421 (PI Bradley Hyman, MD, PhD), P30 AG066509 (PI Thomas Grabowski, MD), P30 AG066514 (PI Mary Sano, PhD), P30 AG066530 (PI Helena Chui, MD), P30 AG066507 (PI Marilyn Albert, PhD), P30 AG066444 (PI David Holtzman, MD), P30 AG066518 (PI Lisa Silbert, MD, MCR), P30 AG066512 (PI Thomas Wisniewski, MD), P30 AG066462 (PI Scott Small, MD), P30 AG072979 (PI David Wolk, MD), P30 AG072972 (PI Charles DeCarli, MD), P30 AG072976 (PI Andrew Saykin, PsyD), P30 AG072975 (PI Julie A. Schneider, MD, MS), P30 AG072978 (PI Ann McKee, MD), P30 AG072977 (PI Robert Vassar, PhD), P30 AG066519 (PI Frank LaFerla, PhD), P30 AG062677 (PI Ronald Petersen, MD, PhD), P30 AG079280 (PI Jessica Langbaum, PhD), P30 AG062422 (PI Gil Rabinovici, MD), P30 AG066511 (PI Allan Levey, MD, PhD), P30 AG072946 (PI Linda Van Eldik, PhD), P30 AG062715 (PI Sanjay Asthana, MD, FRCP), P30 AG072973 (PI Russell Swerdlow, MD), P30 AG066506 (PI Glenn Smith, PhD, ABPP), P30 AG066508 (PI Stephen Strittmatter, MD, PhD), P30 AG066515 (PI Victor Henderson, MD, MS), P30 AG072947 (PI Suzanne Craft, PhD), P30 AG072931 (PI Henry Paulson, MD, PhD), P30 AG066546 (PI Sudha Seshadri, MD), P30 AG086401 (PI Erik Roberson, MD, PhD), P30 AG086404 (PI Gary Rosenberg, MD), P20 AG068082 (PI Angela Jefferson, PhD), P30 AG072958 (PI Heather Whitson, MD), P30 AG072959 (PI James Leverenz, MD).

